# Liwen Ablation versus Modified Morrow Myectomy for Obstructive Hypertrophic Cardiomyopathy: 1-Year Hemodynamic and Conduction Outcomes

**DOI:** 10.64898/2026.09.15.26363181

**Authors:** Jinyu Jiao, Yanli Long, Zhengchun Yu, Qingkun Fan, Ming Liu, Rong Zhou

## Abstract

**Background:** In obstructive hypertrophic cardiomyopathy, surgical myectomy is highly effective but carries a recognized risk of postoperative conduction disturbances. Percutaneous intramyocardial septal radiofrequency ablation (Liwen procedure) is a less invasive alternative, but head-to-head comparisons of longitudinal hemodynamic trajectories and conduction system safety remain scarce, limiting evidence-based treatment selection.

**Methods:** This single-center retrospective cohort included 211 patients treated 2018–2023 (124 Liwen, 87 modified Morrow myectomy). The primary longitudinal analysis included 133 patients with 1-year follow-up and serial data (75 Liwen, 58 modified Morrow myectomy).

Repeated outcomes were analyzed using linear mixed-effects models and generalized estimating equations, adjusted for baseline covariates.

**Results:** Both procedures reduced LVOT gradient, but trajectories differed (interaction *P*=0.002). At 1 year, the covariate-adjusted mean LVOT gradient remained higher after Liwen than after myectomy (16.1 vs 1.3 mmHg, *P*<0.001). However, the adjusted 1-year predicted probability of new conduction abnormalities was markedly lower after Liwen (5% vs 62%, *P*<0.001), with no permanent pacemaker implantations in the Liwen group. Most patients achieved gradient <30 mmHg in both groups (82% vs 96%; nominal *P*=0.308). Liwen had shorter procedure duration (80 vs 185 min) and hospital stay (11 vs 16 days). Serious adverse event rates were similar (8.1% vs 9.2%) but profiles differed.

**Conclusions:** Modified Morrow myectomy achieved more complete LVOT gradient reduction at 1 year, whereas Liwen ablation had fewer conduction abnormalities, shorter procedure duration, and shorter hospital stay. These findings highlight a clinically relevant trade-off between more complete hemodynamic relief and lower conduction-system injury and require prospective confirmation.

**Clinical Perspective:** What Is New? In a single-center retrospective cohort of 133 patients with symptomatic obstructive hypertrophic cardiomyopathy, modified Morrow myectomy achieved more complete 1-year left ventricular outflow tract gradient reduction (residual gradient, 1.3 vs 16.1 mmHg), whereas Liwen ablation was associated with markedly fewer new conduction abnormalities (5% vs 62%) and no permanent pacemaker implantations.

What Are the Clinical Implications? These findings highlight distinct procedural trade-offs and support individualized treatment selection based on anatomy, baseline conduction status, procedural risk, local expertise, and patient preferences; prospective multicenter studies with concurrent enrollment are needed to confirm these comparative associations.

## Introduction

Hypertrophic cardiomyopathy (HCM) is characterized by unexplained thickening of the interventricular septum, which causes dynamic left ventricular outflow tract (LVOT) obstruction in approximately two-thirds of patients, contributing to exertional dyspnea, chest pain, syncope, and adverse outcomes. For symptomatic patients with a resting or provoked gradient ≥50 mmHg despite medical therapy, septal reduction therapy is recommended at experienced centers.^1,2^

Surgical modified Morrow myectomy is the established gold standard. Through open-heart surgery with cardiopulmonary bypass, the surgeon directly resects the hypertrophied septum, immediately enlarging the LVOT and addressing associated subvalvular abnormalities. However, the resection plane lies near the conduction system, resulting in new left bundle branch block (LBBB) in many patients, some requiring permanent pacing.^3,4,5,6^

Percutaneous intramyocardial septal radiofrequency ablation (PIMSRA), the Liwen procedure, is a less invasive alternative. Under echocardiographic guidance, a radiofrequency needle is advanced transapically into the septum, inducing thermal necrosis and progressive remodeling.

Early single-arm studies have reported gradient reduction and symptomatic improvement.^7,8,9^ However, direct comparative evidence against surgical myectomy remains limited, particularly regarding the time course of hemodynamic improvement, mitral valve outcomes, and conduction system effects.^10^

We therefore conducted a retrospective longitudinal cohort study comparing Liwen ablation with modified Morrow myectomy. The primary objective was to evaluate 1-year changes in LVOT obstruction; secondary objectives included septal remodeling, mitral valve function, conduction abnormalities, periprocedural safety, and resource utilization.

## Methods

### Study Design and Population

This single-center retrospective cohort study was conducted at Wuhan Asia Heart Hospital, Wuhan, China. Consecutive patients with symptomatic obstructive hypertrophic cardiomyopathy (oHCM) who underwent Liwen ablation or modified Morrow myectomy between January 2018 and December 2023 were identified. Treatment selection was by multidisciplinary heart team discussion; patients were not randomized. The study was conducted in accordance with the Declaration of Helsinki and approved by the Ethics Committee of Wuhan Asia Heart Hospital (2026-YXKY-P038), with a waiver of informed consent due to retrospective deidentified data.

Exclusion criteria were: hybrid Liwen–modified Morrow myectomy, concomitant valve surgery or coronary bypass, Cox-Maze surgery, prior septal reduction therapy, biventricular outflow tract obstruction, severe pulmonary hypertension, and pre-existing permanent pacemaker. After exclusions, the full safety cohort comprised 211 patients (124 Liwen, 87 myectomy), used for safety and resource utilization analyses. The primary longitudinal cohort included 133 patients with ≥12 months of follow-up and adequate serial imaging and ECG data (<25% missing data for the core efficacy variables at each prespecified time point; 75 in the Liwen group and 58 in the myectomy group). Sample size was determined by consecutive patient availability; no formal prospective power calculation was performed.

Liwen procedures were performed in 2022–2023 (median follow-up 12.4 months, IQR 3.1–20.8), whereas myectomy spanned 2018–2023 (median follow-up 38.5 months, IQR 13.5–60.9). This difference in treatment era and follow-up duration is acknowledged as a potential source of time-related bias.

### Baseline Covariate Definitions

Baseline comorbidities were defined as follows: hypertension (prior diagnosis with active medication, or two preoperative readings ≥140/90 mmHg); coronary artery disease (prior myocardial infarction, coronary stenosis ≥50%, prior revascularization, or angina with objective ischemia); diabetes (prior diagnosis with active treatment or two fasting glucose readings ≥7.0 mmol/L); atrial fibrillation (any documented episode on ECG/Holter). Smoking and alcohol were categorized as ever versus never (ever included current use and former use with ≥6 months of abstinence). Family history referred to first-degree relatives with HCM or sudden cardiac death before age 40.

### Treatment Procedures

Liwen ablation was performed under general anesthesia with echocardiographic guidance. A cooled-tip radiofrequency needle was advanced transapically into the hypertrophied basal septum, and energy was delivered at multiple sites to induce localized coagulative necrosis, avoiding the mitral apparatus and coronary arteries.

Modified Morrow myectomy was performed through median sternotomy with cardiopulmonary bypass. After aortotomy, the hypertrophied basal septum was resected under direct visualization to enlarge the LVOT; subvalvular abnormalities were managed as indicated.

### Follow-Up and Outcome Assessment

Assessments were performed at baseline, immediately post-procedure, and at 1, 6, and 12 months. Transthoracic echocardiography followed American Society of Echocardiography recommendations^11^; maximum interventricular septal thickness (IVSmax), systolic anterior motion (SAM), and mitral regurgitation (MR) severity (none/mild/moderate/severe) were assessed from standard views. Twelve-lead ECGs were acquired at each visit. ECG interval and amplitude parameters were measured by automated algorithms with manual over-read by cardiologists blinded to treatment assignment. New conduction abnormality was defined as the new appearance during follow-up of a conduction block that had been absent on the preprocedural ECG. Conduction blocks included complete LBBB, complete right bundle branch block (RBBB), left anterior fascicular block (LAFB), left posterior fascicular block (LPFB), and second-degree or higher atrioventricular block (AVB).^12^ Peak resting LVOT gradient was calculated by continuous-wave Doppler using the simplified Bernoulli equation.

### Study Outcomes

Primary outcome: adjusted mean peak resting LVOT gradient at 1 year.

Secondary outcomes included: (1) longitudinal change in LVOT gradient (group-by-time interaction); (2) obstruction relief (gradient <30 mmHg); (3) maximum interventricular septal thickness (IVSmax) change; (4) SAM presence; (5) moderate or severe MR; (6) new conduction abnormality; (7) periprocedural serious adverse events and 1-year major clinical events; (8) procedure duration and hospital stay.

Exploratory outcomes included: LVOT gradient <50 mmHg, left ventricular ejection fraction (LVEF) change, chest tightness, syncope, change in aortic regurgitation severity, and ECG parameter changes (heart rate, PR interval, QRS duration, QTc interval [Bazett’s formula], Sokolow-Lyon voltage, maximum ST-segment depression).

The periprocedural period was defined as from procedure initiation to hospital discharge. Serious periprocedural adverse events included death, stroke, pericardial effusion requiring intervention, ventricular septal perforation, mediastinal hematoma, emergency cardiac surgery, and permanent pacemaker implantation.

## Statistical Analysis

Continuous variables are presented as mean ± SD or median (IQR); categorical variables as *n* (%). Between-group comparisons used t-tests/Mann-Whitney U tests and chi-square/Fisher exact tests, as appropriate.

Repeated continuous outcomes were analyzed using linear mixed-effects models (LMMs)^13^ with a first-order autoregressive (AR(1)) correlation structure (selected by AIC/BIC comparison; Supplemental Table S1). LVOT gradient was square-root transformed for skewness. Fixed effects included group, time, group-by-time interaction, and prespecified covariates (age, sex, baseline ln(NT-proBNP), baseline LVOT gradient, baseline IVSmax, baseline LVEF, baseline NYHA class II, baseline mild aortic regurgitation, and baseline outcome value). Collinearity and sparse-cell patterns were assessed before model fitting (Supplemental Table S2). Preoperative flow velocity was excluded because of near-perfect collinearity with gradient (VIF > 20). For NYHA functional class, dummy variables were initially constructed with Class I as the reference (Classes II, III, and IV); Class III was excluded due to collinearity with Class II (VIF = 5.66), and Class IV was omitted because no patient in the Liwen group had Class IV symptoms (*n* = 0), leaving only the Class II dummy variable (VIF = 5.62). For aortic regurgitation severity, moderate or severe aortic regurgitation was too sparse to estimate reliably (0 patients in the Morrow group and 1 in the Liwen group); accordingly, aortic regurgitation was dichotomized as mild versus none, and the mild aortic regurgitation dummy variable was retained. Repeated binary outcomes were analyzed using generalized estimating equations (GEEs)^14^ with binomial distribution, logit link, AR(1) working correlation, and robust standard errors.

All prespecified between-group comparisons at 1 year for the 5 secondary efficacy outcomes (obstruction relief, septal thickness, SAM, moderate or severe MR, and new conduction abnormality) were adjusted using the Bonferroni method, with a corrected significance threshold of *P*<0.01 (0.05/5). The primary outcome (LVOT gradient) was tested at the conventional *P*<0.05 level without multiplicity adjustment. Safety outcomes (periprocedural serious adverse events, 1-year major clinical events), resource utilization outcomes (procedure duration, hospital stay), and exploratory outcomes (gradient <50 mmHg, LVEF, chest tightness, ECG parameters) were reported with nominal *P* values without Bonferroni correction. Syncope and aortic regurgitation were analyzed using Fisher exact tests due to very low event counts. No formal pairwise testing was performed at earlier time points; interaction P values are reported for all time points. No subgroup analyses by baseline characteristics were prespecified or performed.

Models were fitted under the missing-at-random (MAR) assumption using all available observations. Multiple imputation was not performed; robustness was assessed via sensitivity analyses, including an expanded dataset of 198 patients with ≥2 serial assessments, alternative covariance structures, alternative gradient transformation, alternative NYHA modeling, exclusion of an influential observation, and stabilized inverse probability of treatment weighting (IPTW)^15^ (Supplemental Tables S3–S10). After stabilized IPTW, most baseline covariates achieved weighted SMD<0.1; however, NT-proBNP (SMD=−0.20) and baseline LVOT gradient (SMD=−0.17) showed residual imbalance and were interpreted with caution (Supplemental Table S7). For ECG parameters (QRS, QTc, Sokolow-Lyon voltage, maximum ST-segment depression), substantial differential missingness occurred in the myectomy group due to post-procedural LBBB rendering measurements unreliable; these results are presented as exploratory and interpreted with caution.

All tests were two-sided. Analyses were performed in SPSS 25.0; graphs were generated in GraphPad Prism 10.0. This study was reported in accordance with the Strengthening the Reporting of Observational Studies in Epidemiology (STROBE) recommendations.^16^

## Results

### Study Cohort and Baseline Characteristics

The full safety cohort included 211 patients (124 Liwen, 87 myectomy), and the primary longitudinal cohort included 133 patients (75 Liwen, 58 myectomy) (Figure 1). Age and sex were similar between groups. However, the myectomy group had more severe baseline disease: higher LVOT gradient (median 83.5 vs 68.0 mmHg, *P*=0.002), greater IVSmax (23 vs 21 mm, *P*=0.032), lower LVEF (56% vs 60%, *P*<0.001), higher NT-proBNP (1358.5 vs 557.2 pg/mL, *P*<0.001), and more advanced NYHA class (*P*<0.001) (Table 1). All patients in the myectomy group and 96% in the Liwen group had SAM at baseline. Baseline syncope was more common in the myectomy group (36.2% vs 21.3%; *P*=0.058), a difference that did not reach statistical significance. These differences were accounted for in adjusted analyses.

**Figure 1.**
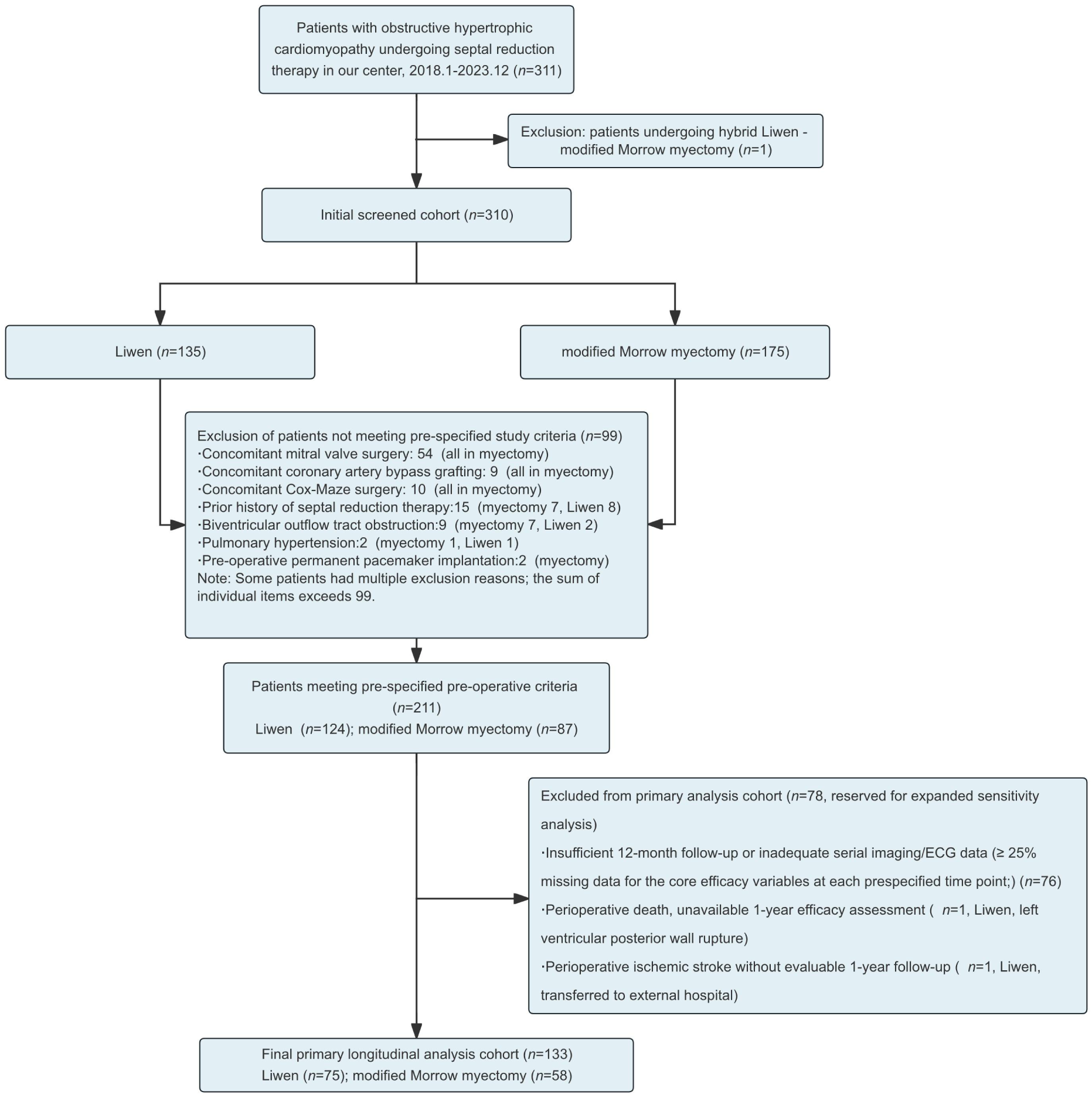
Study flow diagram. Of 311 consecutive patients screened (2018–2023), 1 underwent a hybrid procedure and was excluded, leaving 310. Of these, 99 were excluded for prespecified criteria, leaving 211 in the full safety cohort (124 Liwen, 87 modified Morrow myectomy). Of the 211, 78 were excluded from the primary longitudinal analysis due to <12 months follow-up or <25% missing core efficacy data, leaving 133 (75 Liwen, 58 modified Morrow myectomy).

**Table 1.** Baseline Characteristics of Patients Undergoing Liwen Ablation vs Modified Morrow Myectomy (Primary longitudinal cohort, n=133)

| Characteristic | Liwen group<br>(n=75) | Modified Morrow group<br>(n=58) | P value |
| --- | --- | --- | --- |
| Demographic characteristics |  |  |  |
| Age, y# | 53.0 (42.5, 65.0) | 52.5 (44.0, 58.0) | 0.605 |
| Female, n (%)# | 36 (48.0) | 32 (55.2) | 0.412 |
| Smoking history, n (%) | 21 (28.0) | 20 (34.5) | 0.422 |
| Alcohol consumption, n (%) | 8 (10.7) | 10 (17.2) | 0.272 |
| Family history of HCM, n (%) | 10 (13.3) | 9 (15.5) | 0.721 |
| Comorbidities |  |  |  |
| Hypertension, n (%) | 38 (50.7) | 22 (37.9) | 0.143 |
| Diabetes mellitus, n (%) | 9 (12.0) | 5 (8.6) | 0.529 |
| Coronary artery disease, n (%) | 16 (21.3) | 6 (10.3) | 0.091 |
| Atrial fibrillation history, n (%) | 1 (1.3) | 3 (5.2) | 0.317 |
| Clinical symptoms and cardiac function |  |  |  |
| Preoperative chest tightness, n (%) | 63 (84.0) | 52 (89.7) | 0.344 |
| Preoperative syncope, n (%) | 16 (21.3) | 21 (36.2) | 0.058 |
| Systolic anterior motion (SAM), n (%) | 72 (96.0) | 58 (100) | 0.257 |
| NYHA functional class |  |  | <0.001 |
| Class I, n (%) | 4 (5.3) | 2 (3.4) |  |
| Class II, n (%)# | 59 (78.7) | 30 (51.7) |  |
| Class III, n (%) | 12 (16.0) | 24 (41.4) |  |
| Class IV, n (%) | 0 (0) | 2 (3.4) |  |
| Echocardiographic parameters |  |  |  |
| Preoperative LVOT gradient, mmHg# | 68.0 (38.0, 88.0) | 83.5 (62.0, 106.0) | 0.002 |
| Preoperative LVOT flow velocity, m/s | 4.1 (3.1, 4.7) | 4.6 (3.9, 5.2) | 0.002 |
| IVSmax mm# | 21 (19, 25) | 23 (21, 26) | 0.032 |
| LVEF, %# | 60.0 (58.0, 65.0) | 56.0 (56.0, 60.0) | <0.001 |
| Mitral regurgitation severity |  |  | 0.644 |
| None, n (%) | 1 (1.3) | 1 (1.7) |  |
| Mild, n (%) | 26 (34.7) | 16 (27.6) |  |
| Moderate, n (%) | 19 (25.3) | 18 (31.0) |  |
| Severe, n (%) | 29 (38.7) | 23 (39.7) |  |
| Aortic regurgitation severity |  |  | 0.029 |
| None, n (%) | 39 (52.0) | 41 (70.7) |  |
| Mild, n (%)# | 35 (46.7) | 17 (29.3) |  |
| Moderate-severe, n (%) | 1 (1.3) | 0 (0) |  |
| Biomarkers |  |  |  |
| NT-proBNP, pg/mL* | 557.2 (277.1, 1104.5) | 1358.5 (574.1, 2836.0) | <0.001 |
| ln(NT-proBNP)# | 6.33 ± 1.29 | 7.19 ± 1.08 | <0.001 |
| Electrocardiographic parameters |  |  |  |
| Sinus rhythm, *n* (%) | 72/74 (97.3) | 56/58 (96.6) | 1.000 |
| Mean heart rate, bpm | 70.3 ± 12.6 [n=65] | 66.8 ± 7.7 [n=51] | 0.063 |
| Preexisting conduction abnormality, *n* (%) | 7/74 (9.5) | 6/58 (10.3) | 1.000 |
| PR interval, ms | 164 (152, 179) [n=65] | 160 (152, 182) [n=51] | 0.747 |
| QRS duration, ms | 106 (99, 113) [n=65] | 104 (96, 110) [n=45] | 0.335 |
| QTc interval, ms | 452.97 ± 24.59 [n=65] | 446.37 ± 23.59 [n=43] | 0.168 |
| Sokolow-Lyon voltage, mV | 3.83 (2.88, 4.66) [n=65] | 3.76 (3.20, 5.15) [n=41] | 0.726 |
| Maximum ST-segment depression, mV | 0.12 (0.08, 0.16) [n=45] | 0.13 (0.06, 0.18) [n=36] | 0.970 |

Data are median (IQR) or mean ± SD for continuous variables, and *n* (%) for categorical variables. #Indicates variables included in multivariable-adjusted models. \*NT-proBNP was natural-log transformed for modeling. HCM = hypertrophic cardiomyopathy; LVOT = left ventricular outflow tract; IVSmax = maximum interventricular septal thickness; LVEF = left ventricular ejection fraction; NYHA = New York Heart Association; NT-proBNP = N-terminal pro-B-type natriuretic peptide; SAM = systolic anterior motion; IQR = interquartile range; bpm = beats per minute.

### Periprocedural Safety and Resource Utilization

Based on the full safety cohort (*n*=211), Liwen ablation had a shorter procedure duration (median 80 vs 185 min, *P*<0.001) and shorter hospital stay than myectomy (median 11 vs 16 days, *P*<0.001). At least one serious periprocedural adverse event occurred in 8.1% (Liwen) and 9.2% (myectomy) (*P*=0.772 by Fisher exact test), but event profiles differed qualitatively (Table 2). In the Liwen group, 8 patients (6.5%) required pericardial drainage for effusion, 1 (0.8%) died from left ventricular posterior wall rupture, and 1 (0.8%) had ischemic stroke. In the myectomy group, 2 (2.3%) had ventricular septal perforation, 2 (2.3%) had mediastinal hematoma, and 4 (4.6%) required permanent pacemaker implantation. No patient in the Liwen group required pacing.

**Table 2.**
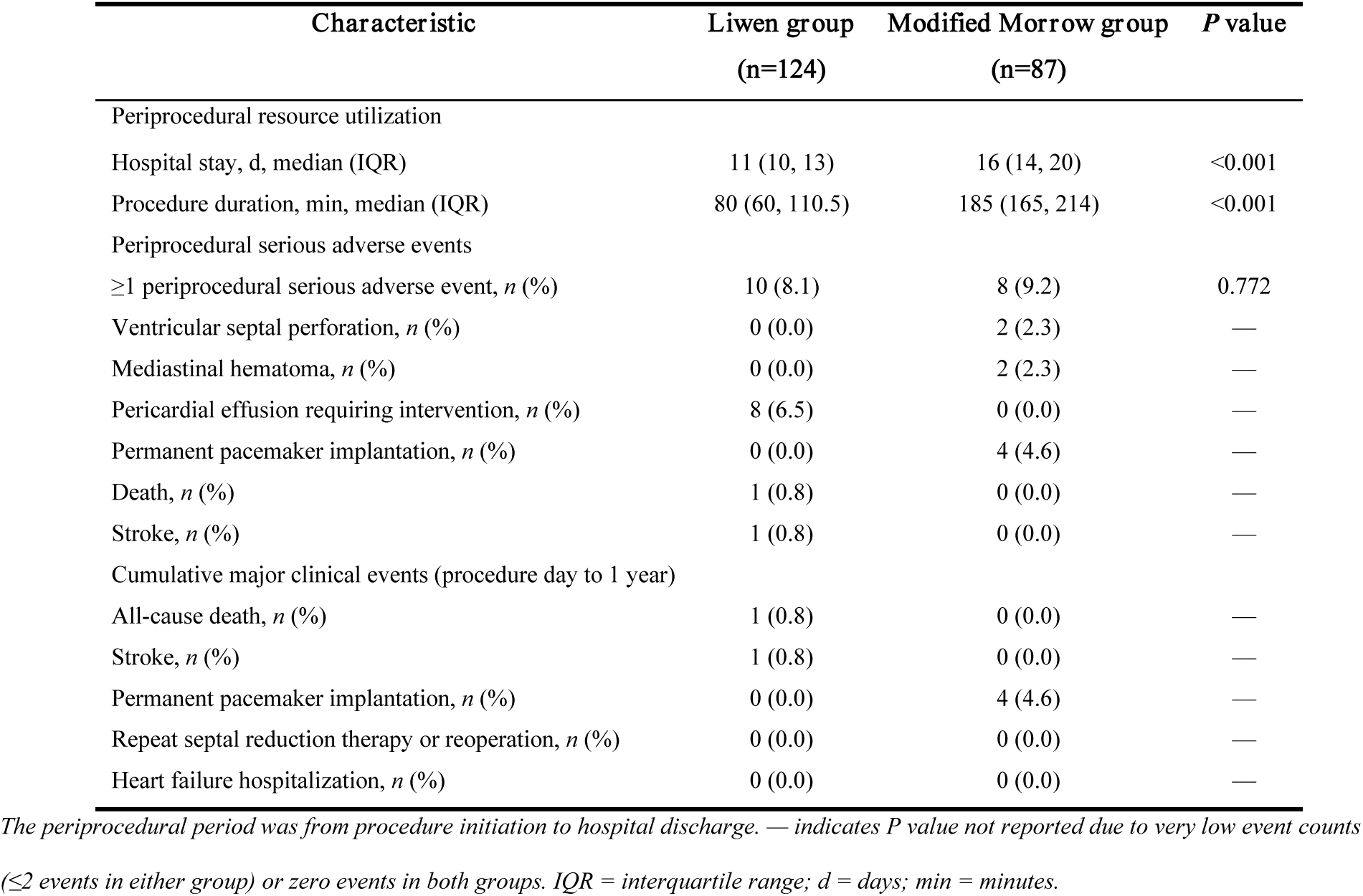
Periprocedural Safety and 1-Year Cumulative Major Clinical Events (Full Safety Cohort, n=211)

Among the primary longitudinal myectomy cohort (*n*=58), 3 patients (5.2%) required permanent pacemaker implantation, whereas 4 of 87 patients (4.6%) in the full safety myectomy cohort required permanent pacemaker implantation. No additional deaths, strokes, repeat interventions, or heart failure hospitalizations occurred after discharge through 1 year; all major clinical events were periprocedural.

### Left Ventricular Outflow Tract Gradient

Both procedures markedly reduced resting LVOT gradient, but their trajectories differed significantly (interaction *P*=0.002) (Table 3, Figure 2A). Myectomy achieved immediate and near-complete reduction (2.0 mmHg immediately post-procedure, 1.3 mmHg at 1 year), whereas Liwen ablation showed gradual decline (32.4 mmHg immediately, decreasing to 16.1 mmHg at 1 year). At 1 year, the adjusted mean LVOT gradient was significantly higher after Liwen ablation than after myectomy, with a between-group difference of 14.8 mmHg (95% CI, 9.7–19.9 mmHg, approximated by the delta method; covariate-adjusted *P*<0.001 from the LMM).

**Table 3.**
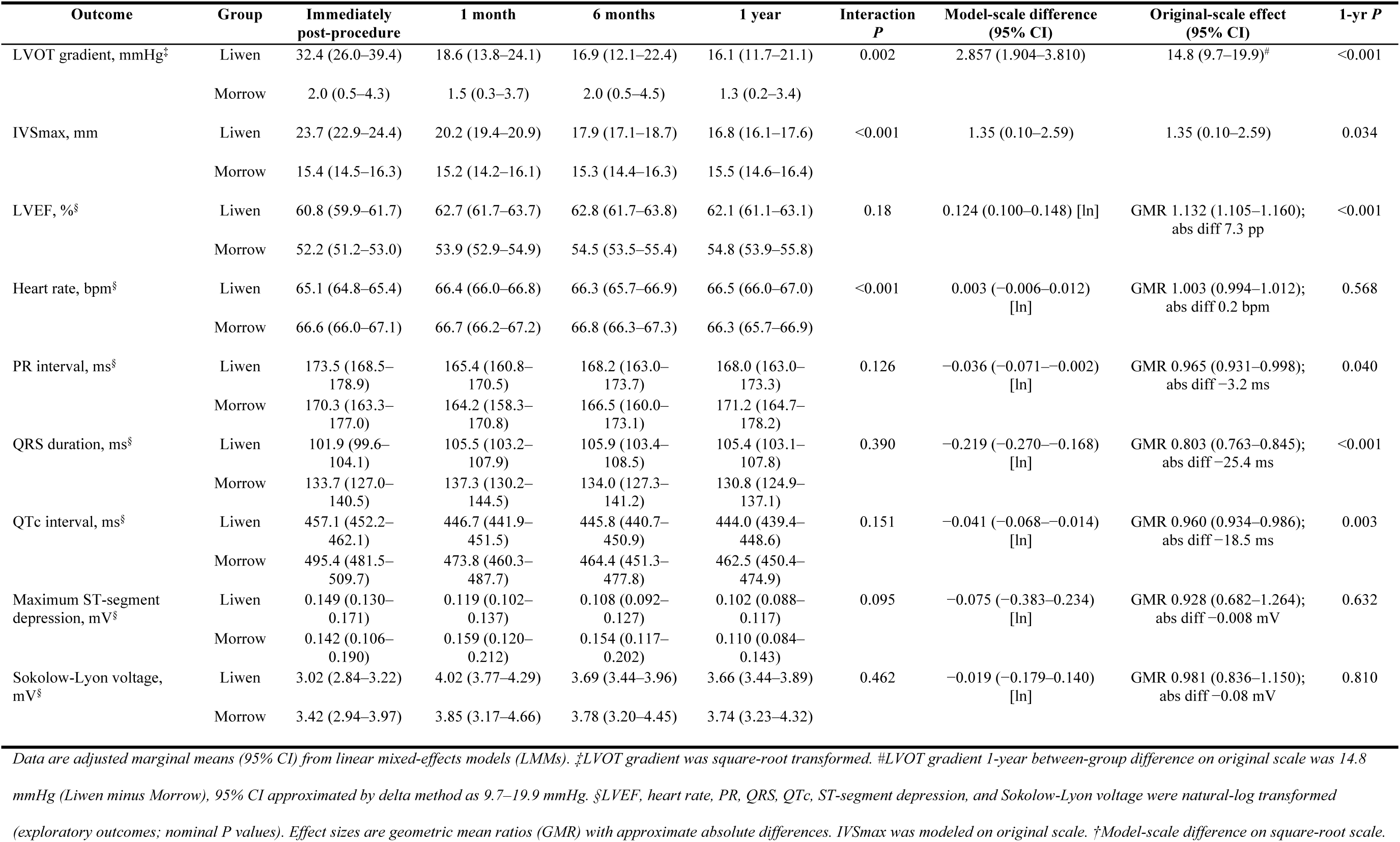
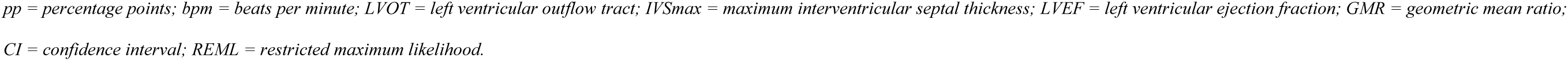
Longitudinal Trajectories of Continuous Outcomes: Adjusted Marginal Means From Linear Mixed-Effects Models (Primary longitudinal cohort, n=133)

**Figure 2.**
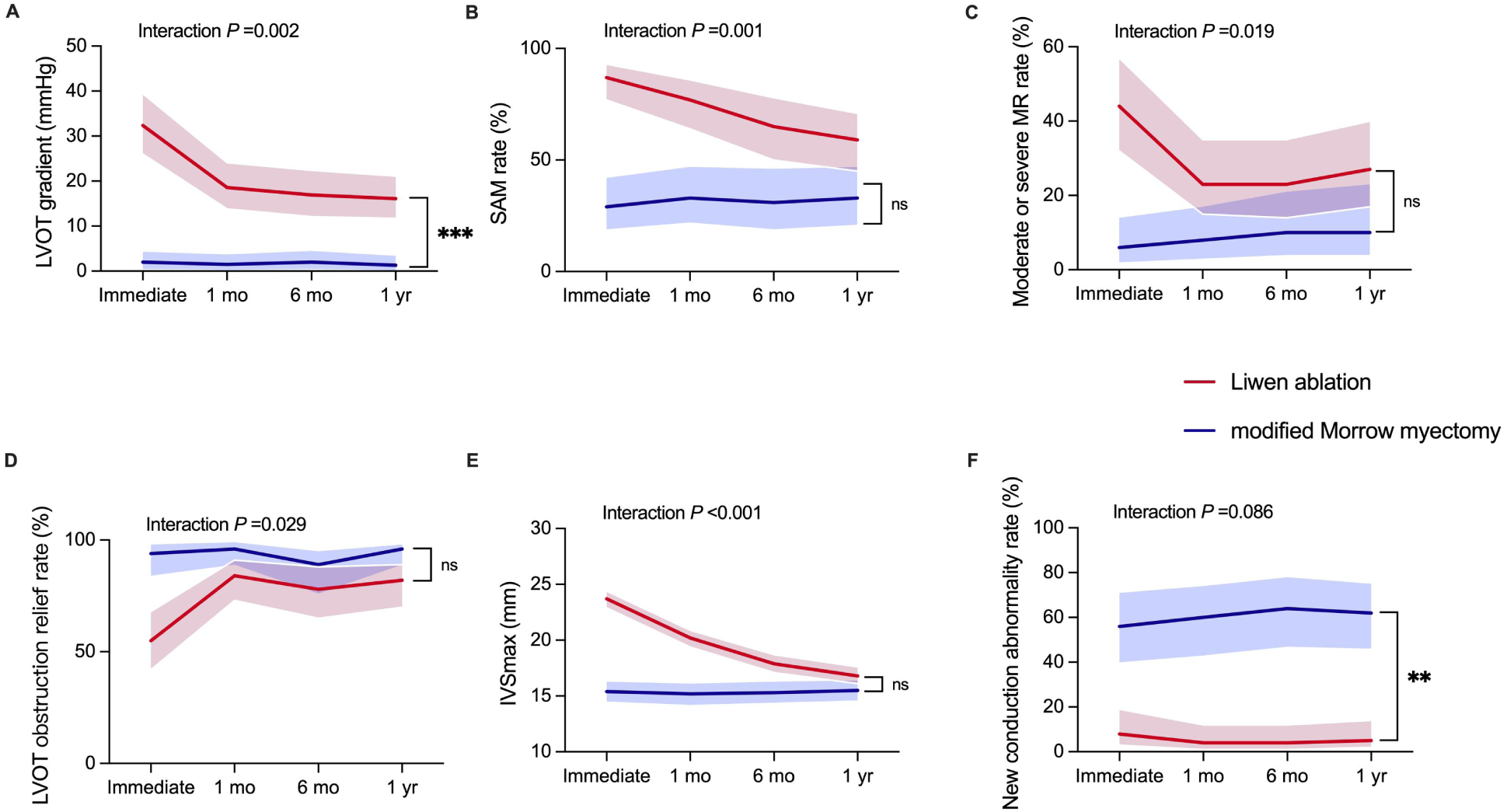
Longitudinal trajectories of key efficacy and safety outcomes after Liwen ablation vs modified Morrow myectomy. (A) LVOT gradient; (B) SAM rate; (C) moderate or severe MR rate; (D) LVOT obstruction relief rate (gradient <30 mmHg); (E) IVSmax; (F) new conduction abnormality rate. Continuous outcomes (A, E): adjusted marginal means with 95% CIs from LMMs; binary outcomes (B–D, F): adjusted predicted probabilities with 95% CIs from GEEs. All models adjusted for prespecified baseline covariates. Interaction P values shown in each panel; brackets indicate 1-year between-group comparisons. Primary outcome (LVOT gradient) tested at P<0.05 without multiplicity adjustment (*P<0.05; **P<0.01; ***P<0.001). Five secondary efficacy outcomes were Bonferroni-corrected (threshold P<0.01; *P<0.01; **P<0.001; ns, not significant). Panel F: 1-year between-group difference was −56 percentage points (95% CI −84 to −29; P<0.001).

The proportion of patients achieving an LVOT gradient <30 mmHg increased in both groups. At 1 year, this was 82% in the Liwen group and 96% in the myectomy group; the difference did not meet the Bonferroni-adjusted threshold (nominal *P*=0.308) (Table 4, Figure 2D).

**Table 4.**
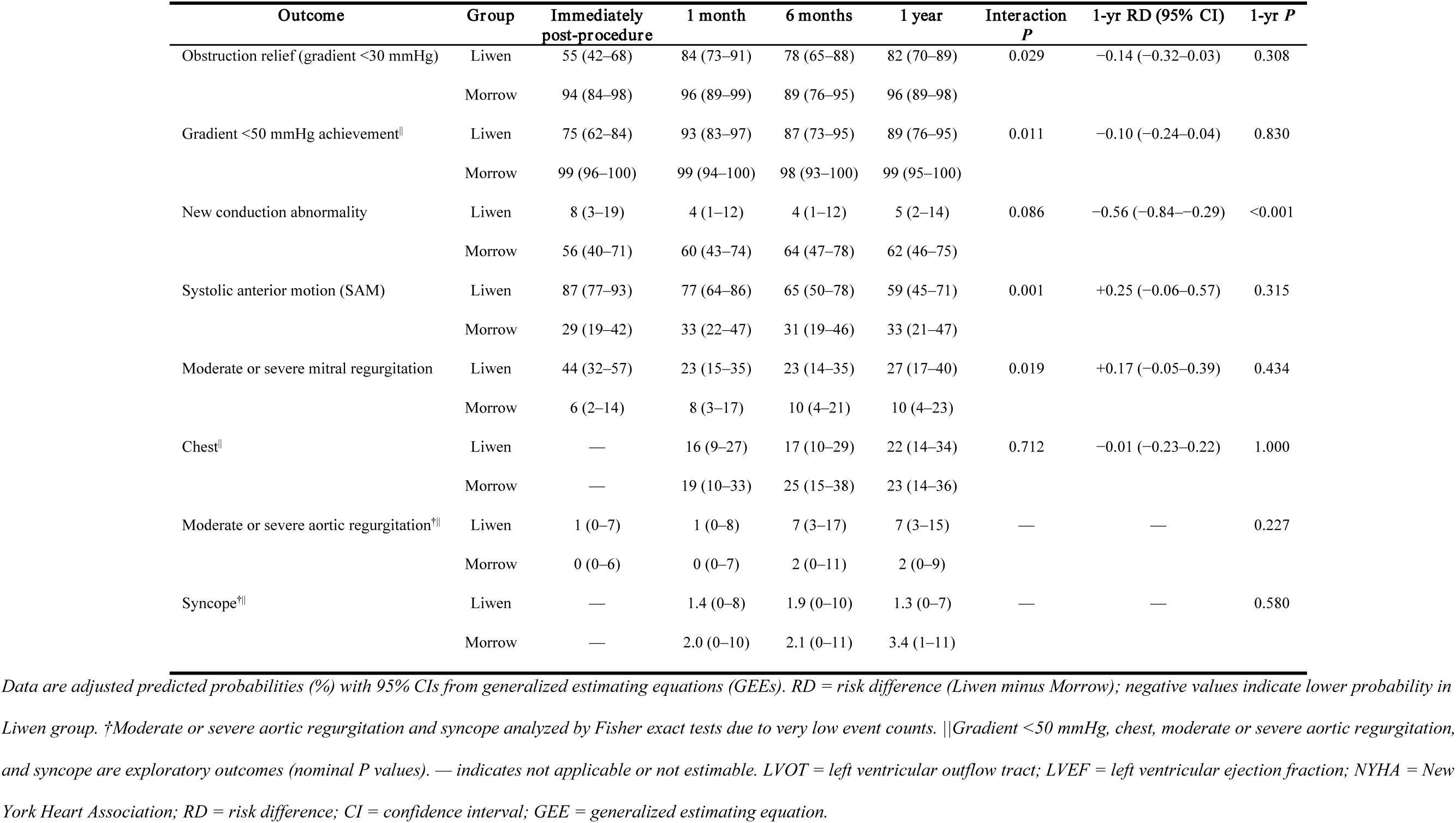
Longitudinal Trajectories of Binary Outcomes: Adjusted Predicted Probabilities From Generalized Estimating Equations (Primary longitudinal cohort, n=133)

### Septal Remodeling and Mitral Valve Outcomes

Septal thickness trajectories differed significantly (interaction *P*<0.001): myectomy caused immediate and stable thinning (15.4 mm immediately, 15.5 mm at 1 year), whereas Liwen ablation showed progressive remodeling, with an apparent early increase to 23.7 mm immediately post-procedure, likely reflecting acute procedural edema and inflammation, followed by gradual thinning to 16.8 mm at 1 year (Table 3, Figure 2E). At 1 year, adjusted septal thickness was slightly greater after Liwen ablation (16.8 vs 15.5 mm, *P*=0.034), but this did not meet the Bonferroni-adjusted threshold (*P*<0.01), and the absolute difference was small (adjusted difference, 1.35 mm [95% CI, 0.10–2.59]), which is of uncertain clinical significance.

SAM prevalence decreased in both groups with differing trajectories (interaction *P*=0.001) (Table 4, Figure 2B). At 1 year, SAM persisted in 59% of the Liwen group and 33% of the myectomy group, which did not meet the Bonferroni-adjusted threshold (nominal *P*=0.315). Moderate or severe MR also decreased with differing trajectories (interaction *P*=0.019) (Table 4, Figure 2C); at 1 year, prevalence was 27% vs 10% (*P*=0.434; did not meet the Bonferroni-adjusted threshold, *P*<0.01).

### Conduction Outcomes

Newly detected postprocedural conduction abnormalities were substantially less frequent after Liwen ablation. At 1 year, the adjusted predicted probability was 5% in the Liwen group and 62% in the myectomy group, corresponding to a risk difference of −56 percentage points (95% CI, −84 to −29; *P*<0.001) (Table 4, Figure 2F). The group-by-time interaction was not significant (*P*=0.086), suggesting no strong evidence that the between-group contrast changed materially over time. Importantly, the 1-year estimate represents predicted prevalence rather than cumulative incidence, because some early conduction abnormalities may be transient.

Among the 37 patients with new conduction abnormalities in the primary longitudinal myectomy cohort (*n*=58), most had complete LBBB (30 of 37, 81.1%). Additional patterns included isolated LAFB (3 of 37, 8.1%), combined RBBB and LAFB (1 of 37, 2.7%), and high-grade atrioventricular block requiring permanent pacemaker implantation (3 of 37, 8.1%, and 5.2% of the entire myectomy cohort). In the Liwen group, 4 of 75 patients (5.3%) developed new conduction abnormalities by 1 year, all of which were complete RBBB; none required permanent pacemaker implantation. This crude observed rate is consistent with the GEE-adjusted predicted probability of 5%.

QRS duration and QTc interval were lower 1 year after Liwen ablation (Table 3), but these exploratory results should be interpreted with caution due to differential non-random missingness in the myectomy group after LBBB development (only 47 and 45 observations, respectively, vs 227 in Liwen), violating the MAR assumption. Similar limitations apply to Sokolow-Lyon voltage and ST-segment depression.

### Exploratory Outcomes

In exploratory analyses, LVOT gradient <50 mmHg at 1 year was achieved in 89% of Liwen patients and 99% of myectomy patients (nominal *P*=0.830). LVEF remained lower in the myectomy group at 1 year (54.8% vs 62.1%, nominal *P*<0.001) without a significant interaction effect (nominal *P*=0.18). Heart rate (66.5 vs 66.3 bpm, nominal *P*=0.568) and chest tightness (22% vs 23%, nominal *P*=1.000) were similar at 1 year, whereas PR interval was slightly shorter after Liwen ablation (168.0 vs 171.2 ms, nominal *P*=0.040) (Table 3). Syncope and moderate or severe aortic regurgitation were rare and did not differ significantly (nominal *P*=0.580 and *P*=0.227, respectively, by Fisher exact tests); these outcomes were not included in the Bonferroni correction because of very low event counts (Table 4). All P values in this paragraph are nominal and unadjusted for multiplicity.

## Sensitivity Analyses

The primary finding of different LVOT gradient trajectories remained consistent across all sensitivity analyses (Supplemental Table S5), including alternative covariance structures (CS: *P*<0.001; UN: *P*=0.004), alternative gradient transformation (ln[gradient + 1]: *P*=0.023), alternative NYHA modeling (continuous 1–4; *P*=0.002), and exclusion of an influential observation (*P*<0.05). In the expanded dataset of 198 patients, the interaction remained significant (*P*<0.001), with directionally consistent results for obstruction relief, SAM, MR, and new conduction abnormality; septal thickness showed a consistent direction but did not reach statistical significance in the expanded cohort (Supplemental Table S10). Stabilized IPTW-weighted analyses, including both linear mixed-effects models (LMMs) for the 9 continuous outcomes and generalized estimating equations (GEEs) for the 6 binary outcomes, produced direction-and magnitude-concordant results across all 15 prespecified outcomes (Supplemental Table S8).

## Discussion

This retrospective longitudinal study compared Liwen radiofrequency ablation with modified Morrow myectomy in symptomatic oHCM. Three principal findings emerge. First, both procedures substantially reduced LVOT obstruction, but myectomy achieved more complete and immediate gradient reduction, with a lower residual gradient at 1 year. Second, Liwen ablation was associated with markedly fewer newly detected postprocedural conduction abnormalities and no permanent pacemaker implantations. Third, Liwen ablation had shorter procedure duration and shorter hospital stay. These findings highlight distinct trade-offs rather than establishing one procedure as uniformly superior.^17^

Modified Morrow myectomy achieved an immediate and near-complete reduction in LVOT gradient, whereas Liwen ablation showed a more gradual decline over the 1-year follow-up period (group-by-time interaction *P*=0.002). This difference has a plausible mechanistic basis: myectomy directly resects obstructive septal tissue to produce immediate anatomical and hemodynamic improvement; in contrast, PIMSRA induces thermal injury and coagulative necrosis, followed by resorption of necrotic septal tissue and delayed, progressive remodeling. The gradual hemodynamic trajectory aligns with prior single-arm PIMSRA/Liwen cohorts, which reported meaningful LVOT gradient reduction but lacked a direct surgical comparator.^7,8,9^ Contemporary surgical series have demonstrated immediate gradient relief after modified Morrow myectomy, with outcomes modulated by surgical technique, concomitant intervention, institutional volume, and operator experience.^4,5,6,18^

Although residual gradient remained higher after Liwen ablation (16.1 vs 1.3 mmHg), most patients in both groups achieved clinical relief (gradient <30 mmHg, 82% vs 96%; between-group difference not statistically significant after Bonferroni correction). This nonsignificant difference does not establish equivalence; the study was not powered for noninferiority, and the point estimate favored myectomy with respect to both lower residual gradient and higher likelihood of gradient <30 mmHg. Whether ongoing septal remodeling beyond 1 year could further narrow this gradient difference requires longer-term follow-up, ideally in prospective contemporaneous cohorts with concurrent assessment of hemodynamic, conduction, and clinical outcomes.

The substantially lower rate of newly detected postprocedural conduction abnormalities after Liwen was clinically notable. This finding is directionally consistent with prior Liwen cohorts, in which permanent pacemaker implantation was uncommon or absent.^7,9^ However, cross-study comparisons should be interpreted cautiously because definitions of conduction abnormality, ECG surveillance, and pacing indications are not uniform. In surgical series, postoperative conduction disturbance remains a recognized trade-off of myectomy, especially when the resection plane is close to the conduction system.^3,4,5,6^

Adverse-event profiles differed qualitatively: pericardial effusion has been characteristic of transapical Liwen approach^7,9^ whereas surgical myectomy has been associated with ventricular septal defect, mediastinal bleeding, and conduction disease.^6,18,19^ Because the study was not powered for rare individual events, the similar composite serious-event rate should not be interpreted as equivalent safety.

Liwen ablation was associated with shorter procedure duration and hospital stay, plausibly reflecting the absence of sternotomy and cardiopulmonary bypass.^7,9^ However, because the groups were treated in different eras, these differences may also reflect evolving perioperative pathways and discharge practices; they should be interpreted as associations rather than causal effects.^18^

## Strengths

This study used longitudinal repeated-measures models that leveraged all available serial assessments, providing greater efficiency and trajectory characterization than single time-point comparisons. Safety was assessed in the full eligible cohort rather than complete-case patients, reducing selection bias. Principal findings were consistent across multiple sensitivity analyses, including alternative model specifications, IPTW weighting, and an expanded dataset.

## Limitations

First, this was a nonrandomized, single-center retrospective study. Treatment selection was by clinical judgment, and the myectomy group had more severe baseline disease. Although multivariable adjustment and IPTW addressed measured confounding, residual unmeasured confounding cannot be excluded.

Second, only 133 of 211 eligible patients were in the primary longitudinal analysis. The expanded sensitivity analysis supports consistency, but selection bias related to incomplete follow-up cannot be eliminated. Excluded Liwen patients had thicker septa and higher NT-proBNP levels, suggesting that the complete-case Liwen cohort may have been somewhat less severely ill than the overall Liwen population.

Third, Liwen procedures were performed in 2022–2023, whereas myectomy spanned 2018–2023, introducing potential era effects related to evolving diagnostic standards, perioperative management pathways, operator experience, and discharge practices. Because the treatment eras did not fully overlap, procedural effects and temporal effects cannot be completely disentangled in the present dataset.

Fourth, preprocedural medication exposure was not systematically collected and may have confounded symptom outcomes. Fifth, follow-up was limited to 1 year, and the study was underpowered for rare events. Sixth, only resting echocardiographic measurements were available; exercise-provoked obstruction, functional capacity, quality of life, and cardiac MRI were not assessed. Seventh, ECG parameters had substantial differential non-random missingness in the myectomy group after LBBB, violating the MAR assumption; these results are exploratory only. Finally, single-center results may not generalize to centers with different expertise.

## Conclusions

Modified Morrow myectomy provided more complete and immediate LVOT gradient reduction at 1 year, whereas Liwen ablation was associated with substantially fewer conduction abnormalities, no permanent pacemaker implantations, shorter procedure duration, and shorter hospital stay. Most patients in both groups achieved hemodynamic relief of obstruction. Given the nonrandomized design, baseline imbalance, different treatment eras, and incomplete follow-up, these results represent comparative associations rather than causal effects. Treatment selection should be individualized based on anatomy, conduction status, procedural risk, local expertise, and patient preferences. Prospective multicenter studies with standardized echocardiographic, ECG, cardiac magnetic resonance, functional, and patient-reported outcomes are needed to confirm these associations.

## Acknowledgments

The authors thank the patients and their families. The authors acknowledge the echocardiography laboratory, electrophysiology team, and clinical laboratory at Wuhan Asia Heart Hospital for data acquisition.

The corresponding author (Ming Liu) had full access to all the data in the study and takes responsibility for the integrity of the data and the accuracy of the data analysis. AI assistance statement: Automated writing assistance tools were used for language editing and reference formatting only; all scientific content, data analysis, and interpretation were performed by the authors, who take full responsibility for the accuracy and originality of the work.

## Author Contributions

Jinyu Jiao contributed to conceptualization, data curation, formal analysis, investigation, methodology, and writing of the original draft. Yanli Long contributed to data curation, investigation, and resources. Zhengchun Yu contributed to data curation, investigation, and validation of echocardiographic measurements. Qingkun Fan contributed to data curation and laboratory resources. Ming Liu and Rong Zhou, the co-corresponding authors, contributed to conceptualization, funding acquisition, project administration, supervision, and writing—review and editing. All authors read and approved the final version of the manuscript and agree to be accountable for all aspects of the work.

## Data Availability

The deidentified individual-level data underlying the reported results, the analytic code, and the data dictionary are not publicly posted because of institutional and ethical restrictions related to patient privacy. Qualified researchers may request access from the corresponding author at. Requests should include a methodologically sound proposal and, where applicable, institutional ethics approval and a data-use agreement. The Wuhan Asia Heart Hospital Ethics Committee and the corresponding author will be responsible for reviewing requests and maintaining data availability.

## Non-standard Abbreviations and Acronyms

HCM: hypertrophic cardiomyopathy
oHCM: obstructive hypertrophic cardiomyopathy
LVOT: left ventricular outflow tract
IVSmax: maximum interventricular septal thickness
LVEF: left ventricular ejection fraction
SAM: systolic anterior motion
MR: mitral regurgitation
NYHA: New York Heart Association
PIMSRA: percutaneous intramyocardial septal radiofrequency ablation
LMM: linear mixed-effects model
GEE: generalized estimating equation
IPTW: inverse probability of treatment weighting
SMD: standardized mean difference
AR(1): first-order autoregressive
CS: compound symmetry
UN: unstructured
NT-proBNP: N-terminal pro-B-type natriuretic peptide
LBBB: left bundle branch block
RBBB: right bundle branch block
LAFB: left anterior fascicular block
LPFB: left posterior fascicular block
AVB: atrioventricular block
VIF: variance inflation factor
AIC: Akaike information criterion
BIC: Bayesian information criterion
MAR: missing-at-random
GMR: geometric mean ratio
REML: restricted maximum likelihood

